# Hypnosis and music interventions for pain, anxiety, sleep, and well-being in palliative care: a systematic review and meta-analysis

**DOI:** 10.1101/2022.01.20.22269568

**Authors:** Josiane Bissonnette, Émilie Dumont, Anne-Marie Pinard, Mathieu Landry, Pierre Rainville, David Ogez

## Abstract

**Background:** Maintaining quality of life is a primary goal of palliative care (PC). Complementary interventions can help meet the needs of patients at the end of life.

**Objectives:** This meta-analysis aims to 1) evaluate the feasibility, acceptability, and fidelity of music and hypnosis interventions designed for patients in PC; 2) evaluate the impact of these interventions on pain, anxiety, sleep, and well-being.

**Methods:** Relevant studies were sourced from major databases. We selected both randomised controlled trials (RCT) and studies relying on pre-post design with details of the intervention(s).

**Results:** Four RCT and seven non-randomised pre-post studies met the inclusion criteria. Overall, the feasibility and acceptability of the interventions reached an adequate level of satisfaction. However, only three studies reported using a written protocol. The meta-analysis of RCT indicated a significant decrease in pain with an effect size of −0.42, *p* = .003. The small number of RCT studies did not allow us to quantify the effects for other variables. Analyses of data from pre-post designs indicated a favourable outcome for pain, anxiety, sleep, and well-being.

**Conclusion:** Despite the limited number of studies included in our meta-analysis, hypnosis and music intervention in the context of PC shows promising results in terms of feasibility and acceptability, as well as improvements on pain, anxiety, sleep and well-being. The available studies are insufficient to compare the efficacy across interventions and assess the potential benefits of their combinations. These results underscore the importance of further research on well-described complementary interventions relying on hypnosis and music.

**Key Messages Box:** What was already known?

- There is a need for validated complementary palliative care interventions.
- Music and hypnosis intervention have shown significant effects for pain, anxiety, sleep, and well-being management in many populations.

What are the new findings?

- Hypnosis and music interventions show medium effect size for pain reduction in palliative care.
- The preliminary analysis of pre-post data shown promising results for pain, anxiety, sleep, and well-being in palliative care.

What is their significance?

- The good feasibility and acceptability of these interventions justify their use in clinical settings.
- More RCT studies with manualised interventions are needed to standardise the procedures, determine effect size and allow for a systematic comparison across interventions.

## Introduction

In 2020, 16.8% of the North American and 20.8% of the European Union population were 65 years or older.[1] Hundreds of thousands of elderly people are or will face a life-threatening illness in the short or medium term and will likely require access to palliative care services. According to the World Health Organization,[2] holistic psychological, social, and spiritual care is a priority for patients who no longer respond to curative interventions. Palliative care (PC) is an approach that improves the quality of life of patients and their families facing problems related to an incurable and/or severe disease with a poor prognosis through the prevention and relief of suffering.[2]

Improving the quality of life for terminally ill patients represents a fundamental clinical issue, as emphasized by many international and national organisations. This unique period of life is represents a critical period characterized by a convergence of factors and considered very important by patients and their families.[3] However, the quality of life of the dying person represents an ideal, one which is often very uncertain because of the predominance of pain, anxiety, depression, and suffering experienced at this stage of life.[3]

Complementary approaches are often centred on mind-body techniques, which may help older individuals live better in the final period of their lives. The minds-body techniques target several problems, both from a somatic and psychological point of view. Such techniques include hypnosis[4] and music[5], wherein interventions based on hypnotic suggestions and musical immersion aims to achieve the objectives of palliative care and improve the overall wellbeing of patients.

Hypnosis is a technique that combines suggestion, relaxation, and imagery to induce an altered state of consciousness to modify one’s subjective experiences, reactions, and behaviours in a given situation.[6] Previous meta-analyses of clinical hypnosis have confirmed its effectiveness in addressing pain, anxiety, and well-being. A first meta-analysis (k = 12) showed that hypnosis provided a larger reduction in chronic pain compared with standard care with moderate effect sizes in favour of hypnosis (g = 0.6, p <.05).[7] A second meta-analysis examined the effect of hypnosis on anxiety in cancer patients (k = 20) and showed a large effect size in favour of hypnosis (g = 1.05, p < .01).[8] Finally, a third meta-analysis (*k* =13), showed that hypnosis significantly reduced sleep latency compared to a waitlist control group.[9] These effects need to be tested in older adults and in PC settings.

To our knowledge, no meta-analysis has investigated the effect of hypnosis on pain and anxiety in older people in the context of PC. A recent study evaluated the effect of clinical hypnosis program on chronic pain in women aged 65-87 years living at home.[10] The interventions were administered in the participants’ homes, and consisted of three 15-min sessions over 12 weeks. The results showed a significant decrease in pain perception for “worst pain”, for “medium pain” intensity as well as for “current pain”. Moreover, the benefits observed during the program persisted at the 12-month follow-up.[11] Altogether, previous research demonstrate the benefits of hypnosis on pain, anxiety, and sleep.

Musical interventions, including musicotherapy and music medicine,[12] represent complementary non-pharmacological approaches used to manage pain, anxiety, insomnia and improve well-being. In recent years, studies have investigated the effect of music on physiological components pertaining to emotion, stress, and pain (e.g. dopamine, serotonin, cortisol, heart rate, interleukin-1).[13] Among these, a first meta-analysis showed a mean effect size (SMD = 0.58) in favour of musical interventions to manage pain.[14] A second meta-analysis examined the effects of music on anxiety and showed a significant decrease in self-reported anxiety in nonclinical populations (k = 19) (d = −0.30),[15] and in clinical population (k = 32) (SMD = −0.36).[16] Another meta-analysis (k = 11) in PC patients reported the effect of music therapy in reducing pain, (k = 6) (SMD = −0.44) and anxiety (k = 3) (SMD = −0.68).[17] Lastly, the effects of preferred music on PC participants were explored and revealed positive results in other empirical studies.[18 19] Again, the available results appear to support the development and evaluation of music interventions in some clinical settings.

Interventions combining music and imagery (MI), as well as guided imagery with music (GIM) generally share the same structure as hypnosis interventions.[20] While GIM and MI are quite similar, they nevertheless differ in the verbal exchanges between the participant and the facilitator, wherein the participant describes what he or she is experiencing during GIM, while the facilitator guides the participant to experience the created images even more intensely. [20] A systematic review examined the effect of GIM on healthy adults and on adults with psychological distress or mental or medical diagnoses. It showed positive results particularly on anxiety (k = 3) and quality of life (k = 2).[21]

Music and hypnosis act on pain, anxiety and well-being across multiple mechanisms. Prevailing views on central pain processing [e.g. 22] posit that whereas nociceptive processes may be triggered by sensory activation evoked by an injury, pathology, or inflammation, the experience of pain involves the integration of sensory, affective and cognitive processes across distributed brain networks.[23 24] Hence, sensory experiences or mental states may activate neural networks interfering with nociceptive processes and affecting pain perception through neurocognitive and emotional processes.[25 26]

Music and hypnosis can also alter emotions through different mechanisms. Previous research has already identified several mechanisms concerning the action of music over emotions, including brain stem reflex, rhythmic entrainment, evaluative conditioning, contagion, visual imagery, episodic memory, and musical expectancy.[27] These mechanisms enable the modulation of positive and negative emotions. In this regard, pleasant music has been shown to decrease the level of perceived pain relative to unpleasant one. [28, 29] Music has also been shown to have effects on the level of arousal of individuals and to reduce the magnitude of spinal-motor[30] and autonomic reflexes induced by acute noxious stimuli.[31] On the other hand, hypnosis typically induces relaxation, mental absorption, and a feeling of automaticity. This reflects global changes in activity within brain networks involved in arousal and saliency as well as attention and other central executive functions, including self-monitoring/regulation. The specific brain mechanisms involved in the modulation of experience by hypnotic suggestions further depend on the modality targeted by those suggestions, with significant effects seen in the corresponding brain regions for somatosensory, visual, auditory and motor processes.[32] Depending on the focus of analgesic suggestions, pain modulation by hypnosis may involve changes in brain regions involved in the sensory and/or the affective dimension of the experience[33 34] and those in brain networks promoting mental imagery and self-referential memory processes.[32] Similar to the effects reported in responses to music intervention, the changes in brain activity produced by hypnotic analgesia relate to reductions in autonomic and spinal-motor reflexes, although these effects are generally stronger or observed only in highly hypnotisable individuals (e.g. 35–40). Taken together, mechanistic studies provide strong support for music and hypnosis interventions to effectively harness modulatory brain processes to improve pain management.

Considering the effectiveness of hypnosis and music interventions for the studied population—and for related populations—and the need for complementary interventions in PC, it seems important to draw up a global portrait of these approaches by carrying out a systematic review. The current meta-analysis primarily aims to identify music and hypnosis studies with manualized intervention designed for individuals in PC and to analyse their feasibility, their acceptability, and their fidelity. Next, we also aim to evaluate the impact of music and hypnosis interventions with manualised intervention on pain, anxiety, sleep, and well-being in PC. To this end, the present work compares randomised controlled studies together, and then, to get a broader picture, compares all studies with pre-post data. The outcome will allow us to determine the relevancy of pursuing the development of these interventions.

## Methods

The review has been registered in PROSPERO (CRD-42021236610). No other protocol was published. The search method followed the steps developed by the Preferred Reporting Items for Systematic Reviews and Meta-Analyses (PRISMA) [41] and Cochrane for systematic reviews.[42]

### Eligibility

To consider studies in this systematic review, we established inclusion criteria relating to participants, interventions, study designs, and study variables.

To be included, the study population had to be 18 years or older, have a diagnosis of advanced life-limiting disease, be treated with palliative intent, and not be treated curatively. Interventions had to contain hypnosis and/or music with or without psychoeducational elements. The study had to include a pre-post design in the same participants or a RCT design comparing a group receiving the target intervention to one or more control groups. The intervention protocol also had to be sufficiently detailed to allow for replication and practical implantation in various areas of PC. Studies containing imagery and visualisation were also included, however, mixed interventions combining hypnosis and/or music with other interventions (e.g. massage, acupuncture, writing) in the same intervention were excluded.

Finally, the outcomes assessed included one or more of the following variables: pain, anxiety, sleep, and well-being. Understanding that well-being as a complex variable, we also considered its subcomponents as identified in a meta-analysis, i.e. quality of life, psychological state, life satisfaction, mental well-being, social and spiritual well-being, level of functioning and activities.[43]

### Literature search and article selection

#### Electronic search

A librarian was consulted to develop a literature research strategy. Articles were initially selected using the following search engines: PsychINFO; PubMed; CINAHL Plus; CINAHL; Psychology and Behavioral Sciences Collection and AgeLine from the EBSCO, OVID and EMBASE interfaces, as of January 21, 2021. The following terms were used according to these components: (Interventions) AND (outcomes) AND (clinical settings): (hypnosis or hypnotherapy or hypnoses or hypnotism or hypnotherapy or “guided imagery” or sophrology or music*) AND (pain or anxiety or fear or “fear of dying” or “fear of death”) or (“psychological stress” or distress) or (sleep or insomnia) or well-being or “psychological state” or “quality of life” or “Life satisfaction”) AND (“palliative care” or “end of life care” or “terminal care” or dying or “hospice care”).

We included peer-reviewed scientific articles in English and French in the category “clinical trial”. We excluded theses and dissertations. Details of the search strategy are presented in the online supplemental eTable 1.

**Table 1.**
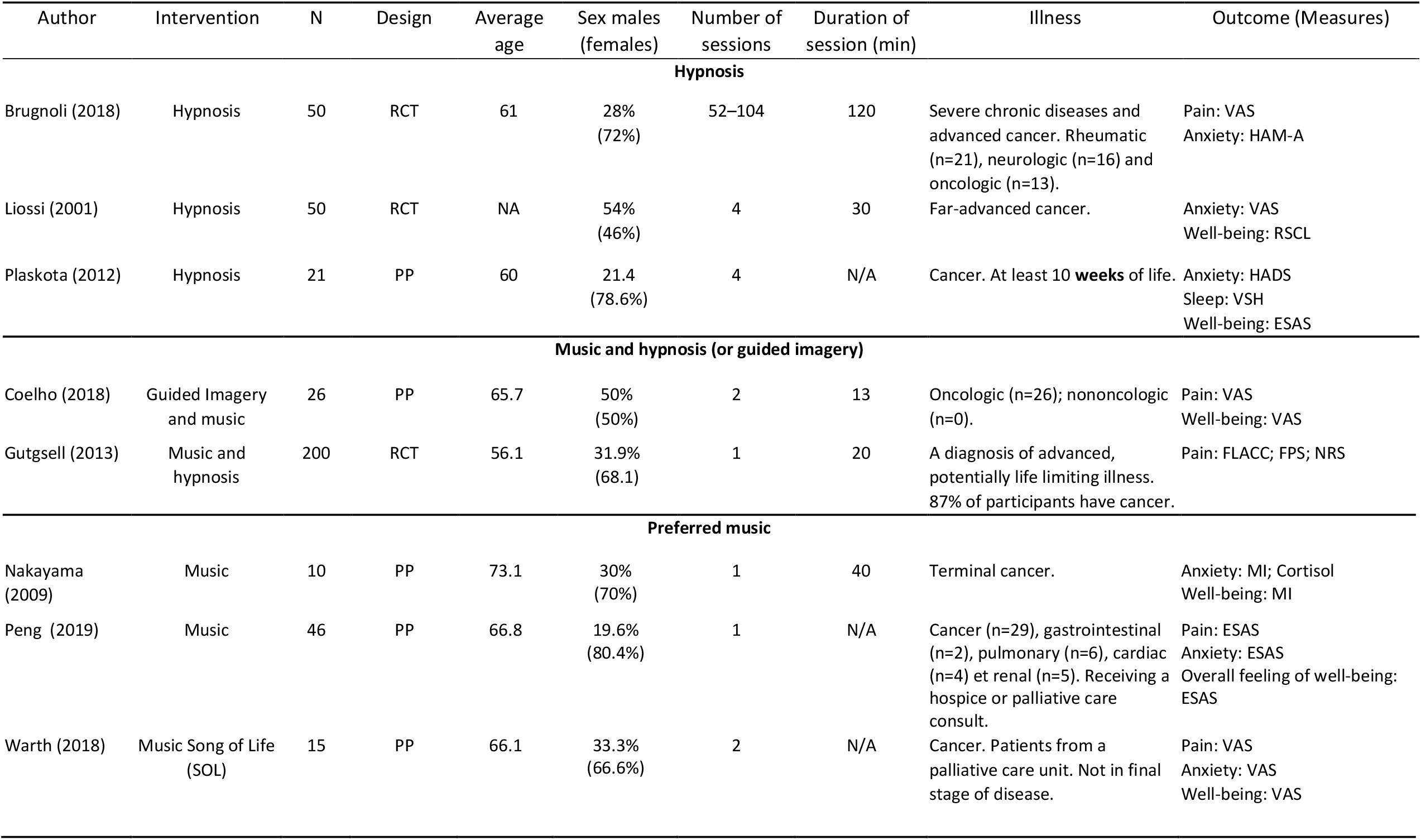
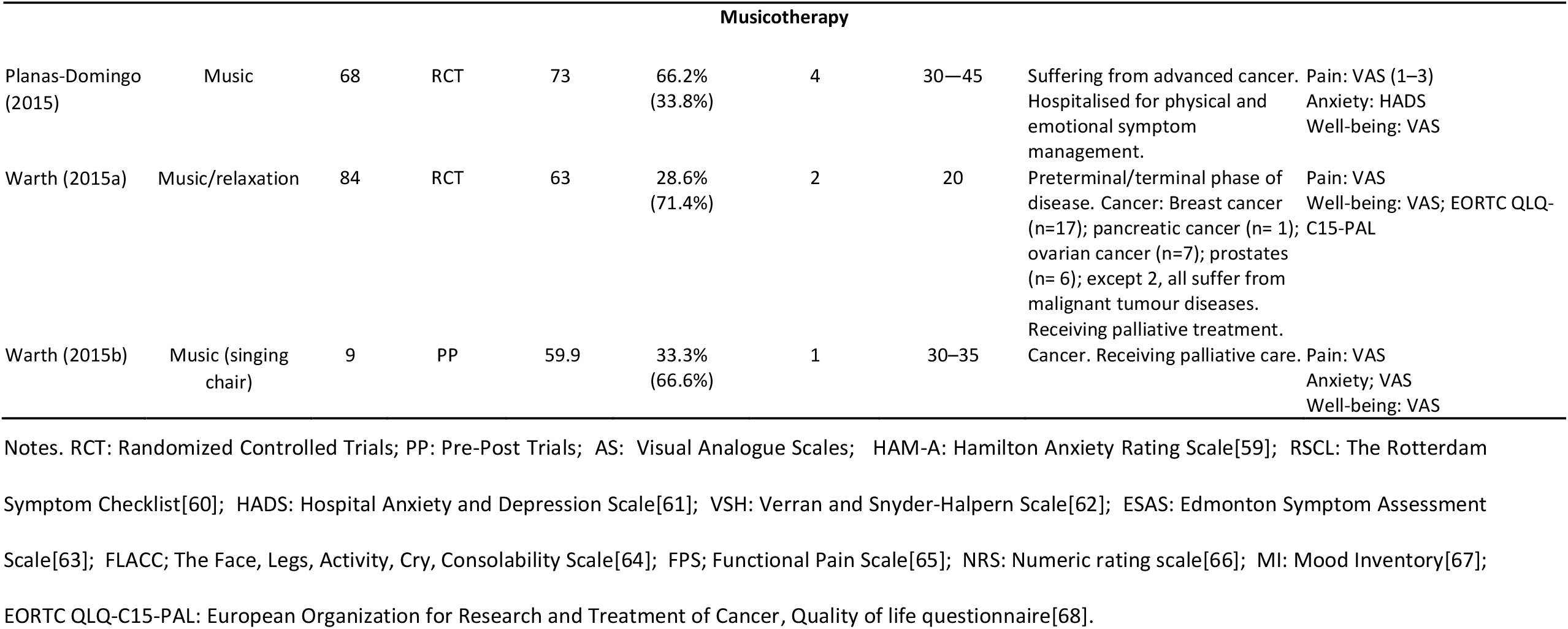
Characteristic of the studies.

#### Data Collection Process

Two authors (JB, ED) independently completed the article selection and data extraction processes according to the steps described below. Disagreements were resolved by consensus.

The authors consolidated the results of the electronic search in Rayyan,[44] a reference management software, and then reviewed the titles and abstracts to exclude irrelevant articles. The articles were then read in their entirety and were included according to the selection criteria. Additional relevant articles from the reference list of every study reports included in the systematic review were then added.

#### Quality assessment of studies

We assessed the quality of the randomised controlled trials (RCT) and pre-post studies using the RoB2 tool by two independent coders.[42] We also assessed the quality of the interventions based on feasibility, acceptability, and fidelity criteria.[45] The acceptability and feasibility of the interventions were assessed in terms of recruitment rates (less than 50% was coded as high risk of bias, more than 85%, as low risk of bias), retention and adherence rates (less than 60% = high risk; more than 85% = low risk), and satisfaction with the approach was evaluated by the synthesis provided by the authors typically from semi-structured interviews they conducted with participants or feedback from health-care professionals. The assessment of the fidelity of interventions was based on the presence of a intervention manual and on the detailed description of the intervention in the article (presence of a intervention manual = low risk, no manual, but detailed intervention = medium risk, neither manual nor detailed intervention = high risk). The presence of a intervention manual should ensure that the intervention has been delivered consistently to users.

#### Data extraction

The following information was extracted from the studies and integrated into an Excel spreadsheet : number of participants recruited, mean age, sex, study design, intervention characteristics (type, duration, number of sessions), clinical characteristics of participants and outcomes (Table 1).

We also extracted information on theoretical models explaining needs and difficulties by patients in PC, on theoretical models explaining impact of intervention, on models of change, and on concrete intervention techniques implemented. Information about acceptability, feasibility, and fidelity of interventions were also noted (online supplemental eTable 2).

**Table 2.**
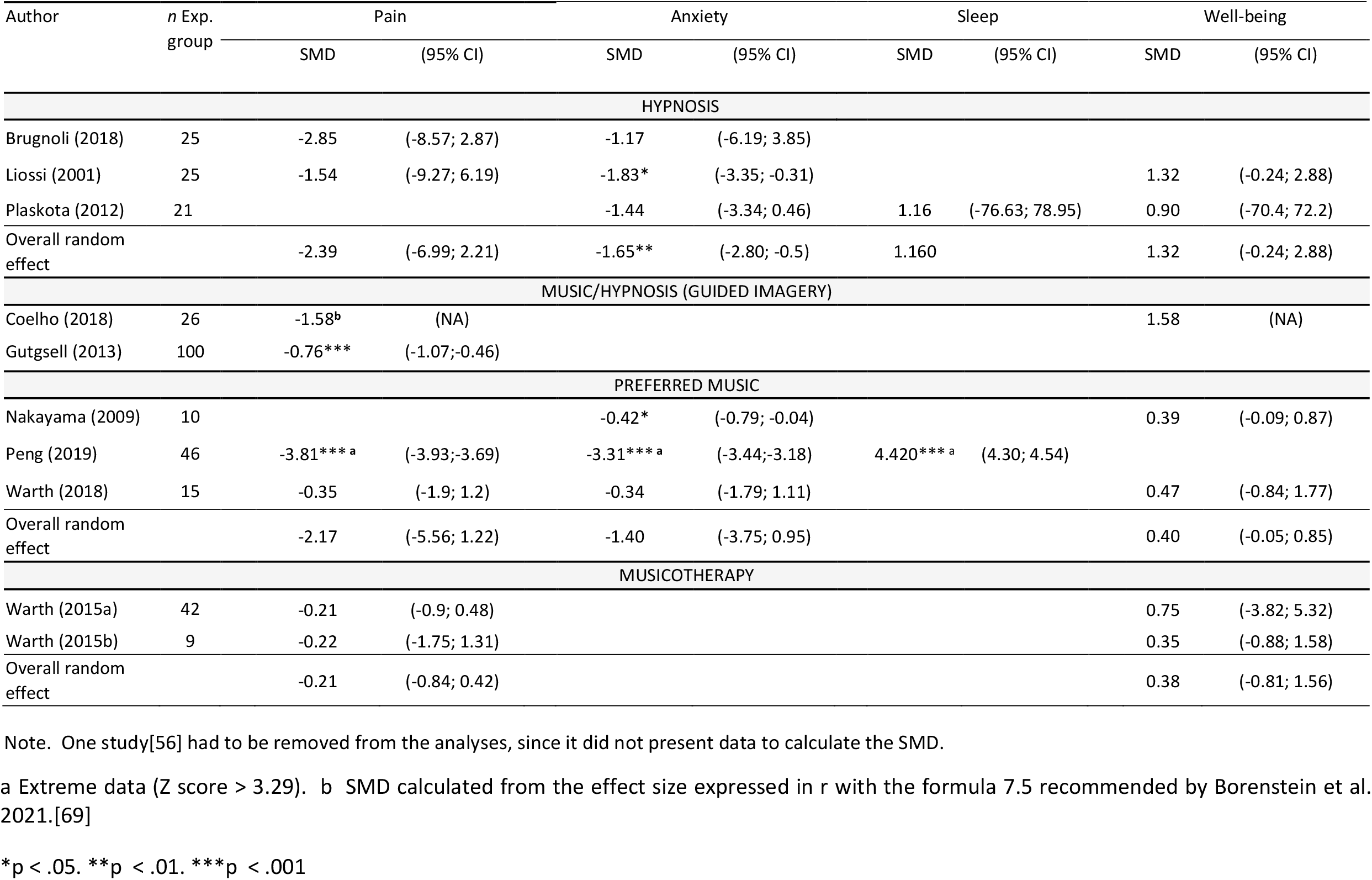
Effect of the interventions on the variables pain, anxiety, sleep and well-being from the data of the experimental group.

| Author | n Exp.<br>group | Pain |  | Anxiety |  | Sleep |  | Well-being |  |
| --- | --- | --- | --- | --- | --- | --- | --- | --- | --- |
|  |  | SMD | (95% CI) | SMD | (95% CI) | SMD | (95% CI) | SMD | (95% CI) |
| HYPNOSIS |  |  |  |  |  |  |  |  |  |
| Brugnoli (2018) | 25 | -2.85 | (-8.57; 2.87) | -1.17 | (-6.19; 3.85) |  |  |  |  |
| Liossi (2001) | 25 | -1.54 | (-9.27; 6.19) | -1.83* | (-3.35; -0.31) |  |  | 1.32 | (-0.24; 2.88) |
| Plaskota (2012) | 21 |  |  | -1.44 | (-3.34; 0.46) | 1.16 | (-76.63; 78.95) | 0.90 | (-70.4; 72.2) |
| Overall random effect |  | -2.39 | (-6.99; 2.21) | -1.65** | (-2.80; -0.5) | 1.160 |  | 1.32 | (-0.24; 2.88) |
| MUSIC/HYPNOSIS (GUIDED IMAGERY) |  |  |  |  |  |  |  |  |  |
| Coelho (2018) | 26 | -1.58<sup>b</sup> | (NA) |  |  |  |  | 1.58 | (NA) |
| Gutgsell (2013) | 100 | -0.76*** | (-1.07;-0.46) |  |  |  |  |  |  |
| PREFERRED MUSIC |  |  |  |  |  |  |  |  |  |
| Nakayama (2009) | 10 |  |  | -0.42* | (-0.79; -0.04) |  |  | 0.39 | (-0.09; 0.87) |
| Peng (2019) | 46 | -3.81***<sup>a</sup> | (-3.93;-3.69) | -3.31***<sup>a</sup> | (-3.44;-3.18) | 4.420***<sup>a</sup> | (4.30; 4.54) |  |  |
| Warth (2018) | 15 | -0.35 | (-1.9; 1.2) | -0.34 | (-1.79; 1.11) |  |  | 0.47 | (-0.84; 1.77) |
| Overall random effect |  | -2.17 | (-5.56; 1.22) | -1.40 | (-3.75; 0.95) |  |  | 0.40 | (-0.05; 0.85) |
| MUSICOTHERAPY |  |  |  |  |  |  |  |  |  |
| Warth (2015a) | 42 | -0.21 | (-0.9; 0.48) |  |  |  |  | 0.75 | (-3.82; 5.32) |
| Warth (2015b) | 9 | -0.22 | (-1.75; 1.31) |  |  |  |  | 0.35 | (-0.88; 1.58) |
| Overall random effect |  | -0.21 | (-0.84; 0.42) |  |  |  |  | 0.38 | (-0.81; 1.56) |

Note. One study[56] had to be removed from the analyses, since it did not present data to calculate the SMD.
a Extreme data (Z score > 3.29). b SMD calculated from the effect size expressed in r with the formula 7.5 recommended by Borenstein et al. 2021.[69]
\*p < .05. \*\*p < .01. \*\*\*p < .001

### Meta-analysis procedures

#### Effect size

The meta-analysis was conducted using Comprehensive Meta-Analysis software version 3.0.[46] There are three basic approaches to comparing the effect size of a set of studies: compare (1) the final values of the control group to those of the experimental group; (2) the pre-test values to the post-test values of the experimental group; and (3) the difference between the pre-test and post-test values of the experimental group to the difference between the pre-test and post-test values of the control group.

In this meta-analysis, we will first present the data from the third approach, as it accounts for the richness of the RCT specifications. To perform the effect size calculations, we used the formula recommended by Morris 2008; (see equation 8, p. 369)[47] with a correction for small sample bias.

Given the small number of studies in the target literature and the fact that half of these studies did not have a control group, we also present the data from the second approach, i.e. comparing the values between the pre-test and post-test for each experimental group of RCT and pre-post designs. This will allow us to compare all selected interventions on the same basis. For the studies that evaluated post-treatment effects at several measurement times, we calculated only the effect size of the first post-intervention data.

To compare the pre-post data, we used the formula of Morris and DeShon (2002)[48] with correction for small sample bias. We calculated the differences in standardised means (SMD) using a random-effect model for all studies considering the clinical heterogeneity of the studies. For each study, a Fisher’s Z-score was calculated to test for the presence of outliers. The homogeneity of effect sizes was examined by the Q-statistic and the I2 value. Publication bias was assessed by the funnel plot, Egger’s regression test and the Duval and Tweedies procedure.[49]

## Results

### Study Selection

A total of 260 articles were identified in the electronic search. After removing duplicates and a careful reading of study titles and abstracts by two coders, we retained 53 studies. We then identified 11 articles that met the inclusion criteria (Figure 1). The references of the selected articles and the reference list of relevant texts were also examined.

**Figure 1.**
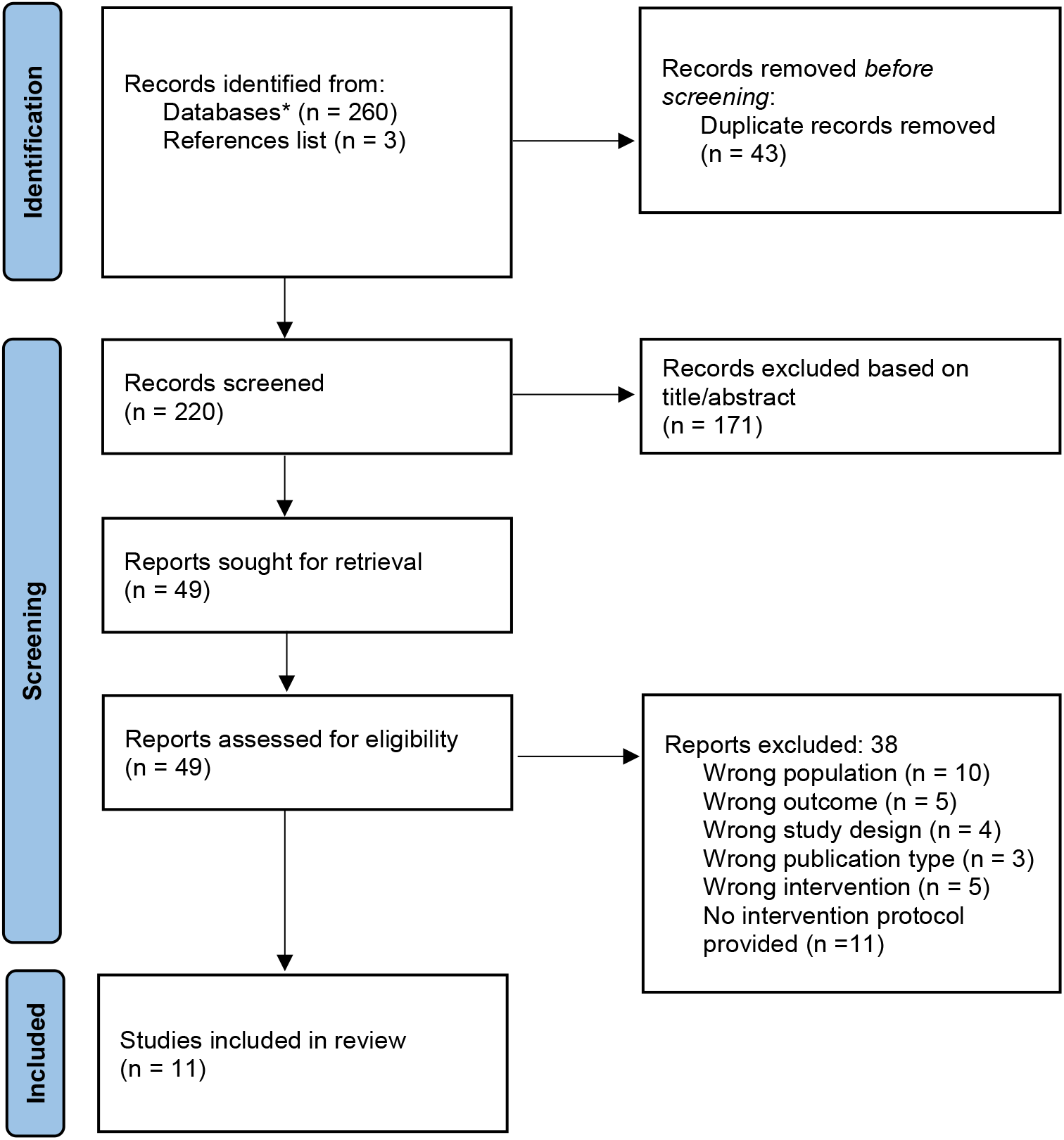
PRISMA Flowchart[41] * Embase, PsychINFO, PubMed, CINAHL Plus, CINAHL, Psychology and Behavioral Sciences Collection, AgeLine.

### Study characteristics and description of the interventions

As presented in Table 1, eleven interventions in hypnosis, hypnosis/music, and music were listed, involving 579 participants with sample sizes ranging from 9 to 200 per study. Three articles evaluated the effect of a hypnosis intervention on at least one of the variables of interest (n = 121), two articles evaluated the effect of a mixed hypnosis and music intervention (n = 226), and six articles evaluated musical interventions (n = 232). The latter were subdivided into two categories: interventions using participants’ preferred music (n = 71) and other musical interventions (n = 161).

Six studies (n = 127) followed a pre-post design[18 19 50–53] and five studies (n = 452) were randomised controlled trials (RCT). Of these five RCT studies, three used a control group with standard pharmacological care[54–56]; one study used a control group with standard pharmacological and psychological care[57] and one study used a control group with standard physiological care with a “meditation” intervention of the same duration as the music group but without musical elements.[58] The number of sessions varied between 1 and 104 and the duration of each varied between 13 and 120 min. The average age of participants per study ranged from 56.1 years to 73.1 years and all participants were in PC, most with cancer (n = 499).

To assess pain, anxiety, sleep, and well-being, we used different measurement tools. Visual analogue scales (VAS) and numerical scales were the most used measurement. One behavioural pain assessment (FLACC) and one biological measure and psychometrics measures were also used (see Table 1).

Regarding the intervention itself, three studies reported following a written protocol and the others detailed their intervention. Information about retention rate, recruitment rate, participant satisfaction, the theoretical models underlying the interventions and the components of each intervention are detailed in the online supplemental eTable 2.

### Effect Sizes

Considering the advantage of RCT over pre-post studies, we will present the results of the meta-analysis of the RCT studies first. We will then present the effect sizes of the changes in the variables between the pre-test and the post-test of all the studies selected. The effect sizes of the pre-post assessment do not take into account the control group of the RCT, however, they do provide insight into the potential impact of the intervention over time.

#### Meta-analysis for data from randomised controlled studies

We performed meta-analyses for data from all RCT[54 55 57 58] for the variable pain including all type of interventions. Since we only had two studies for the anxiety and well-being variables, and no studies for the sleep variable, we did not perform meta-analyses for these three variables. According to the analyses, the overall effect size associated with the interventions is SMD = −0.42 (p = .003; CI = −0.70, −0.14; k = 4). Heterogeneity is negligible (Q = 1.23; p = .74; I2 = 0.00) but the effect appears to be driven by the study of Gutgsell et al.[55] (Figure 2).

**Figure 2.**
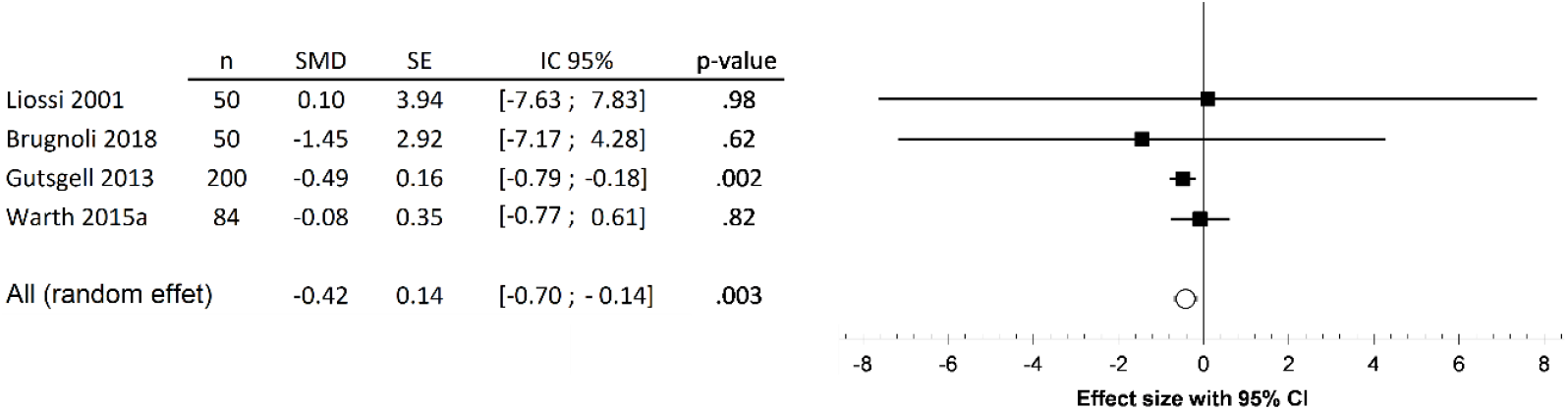
Forest plot – Effect of interventions on pain

The visual analysis of the Funnel Plot does not suggest a publication bias (online supplemental eFigure 1). The Trim and Fill procedure[44] was found not to be significant, showing the absence of publication bias.

#### Pre-Post Effect Sizes per Intervention

In the previous analyses, only four studies were considered. In order to take into account the data from all the interventions selected at the end of the selection procedure, Table 2 presents the pre-post data for each of the studies’ variables.

The data retrieved from the hypnosis interventions showed large effect sizes for the variables pain, anxiety, sleep, and well-being. The overall effect size was significant for global anxiety (SMD = −1.65, p <.01).

For the music/hypnosis interventions, we found a large effect size for pain reduction. This effect was significant for the Gutgsell et al. study[55] (SMD = −0.76, p <.001). Anxiety, sleep, and well-being were not evaluated for this type of intervention.

Effect sizes showed large variations between studies using preferred music, with the study of Peng et al. (2019) presenting extreme data with SMD> 3.3.[18] For the two other studies, the effects are low-moderate for pain, anxiety, and well-being.[19 53] In all cases, the preferred music acts on the four measured variables expectedly.

Two studies that have evaluated music therapy interventions report non-significant effects with small effect sizes for the variable pain and small to moderate effect sizes for well-being.[52 58] Anxiety and sleep were not measured in these studies.

### Quality assessment

#### Risk of bias

The quality of the 11 studies selected was assessed using the RoB2 tool.[42] Only RCT studies have been coded for the Domain 1 (risk of bias arising from the randomization process).The pre-post studies were also assessed using this tool, except for Domain 1, which is exclusively relevant for comparison groups. We also evaluated the acceptability, feasibility, and reliability of the interventions. These results are summarised in Figure 3.

**Figure 3.**
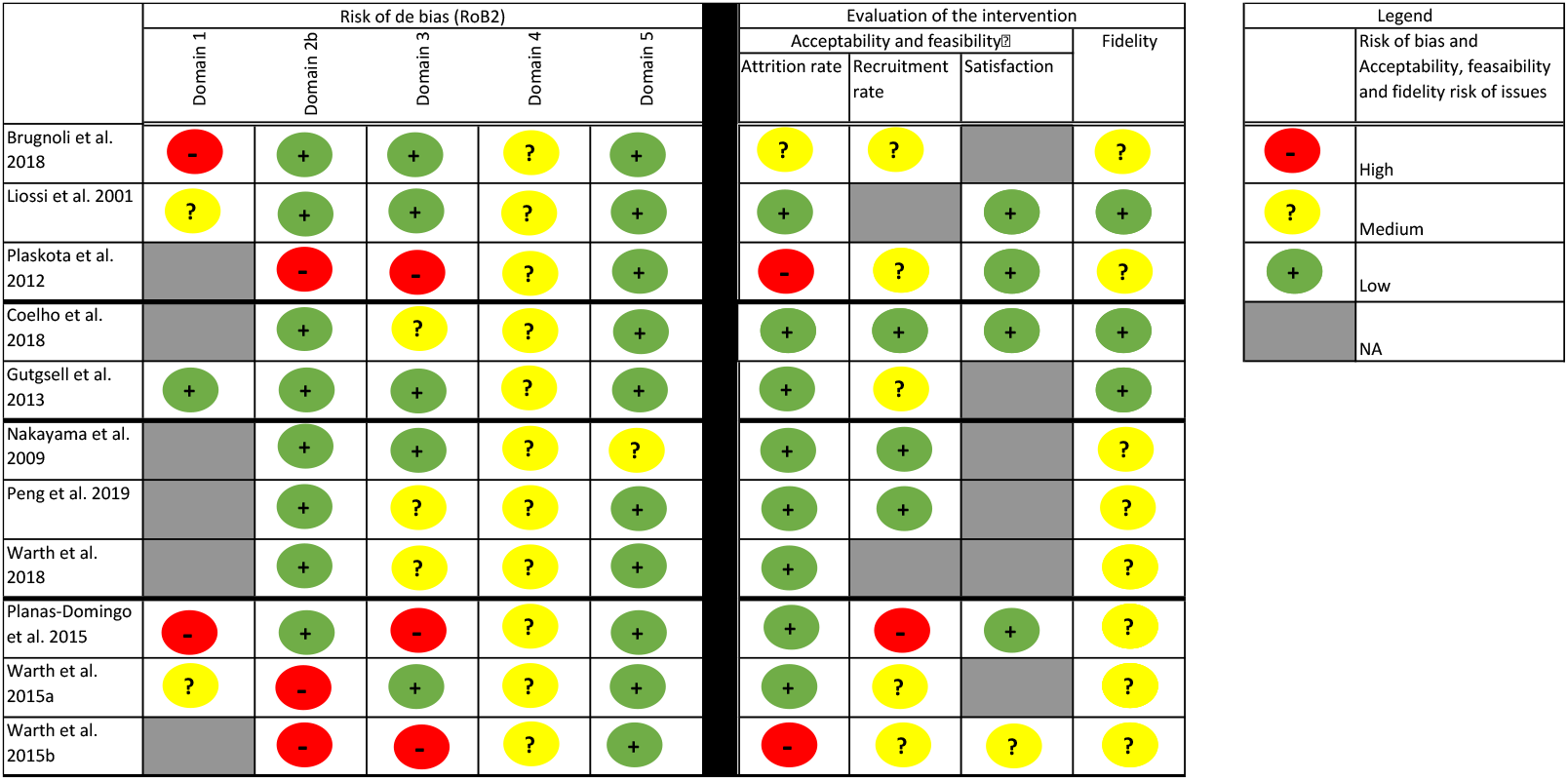
Quality assessment of the studies (RoB2) and evaluation of acceptability, feasibility, and fidelity of interventions. Note. Domain 1: Risk of bias arising from the randomisation process; Domain 2b: Risk of bias due to deviations from the intended interventions (effect of adhering to intervention); Domain 3: Risk of bias due to missing outcome data; Domain 4: Risk of bias in measurement of the outcome; Domain 5: Risk of bias in selection of the reported result.

The randomised and concealed allocation until participants were enrolled and assigned to interventions presents a medium or high risk of bias for almost all studies, except one.[55] All participants and those delivering the interventions were therefore aware of the assigned intervention during the trial but only three studies[51 52 58] present a high risk of bias for Domain 2 due to possible deviations from the intended interventions (effect of adhering to intervention). Non-adherence to the assigned intervention regimen could have also affected outcomes.

The availability of data on attrition rates varied across studies. For two studies, the attrition rate was very high[51 52] and for one study, the attrition rate was not indicated at all.[56] One study showed a very high dropout rate for year 2 but no dropout rate for year 1[54]. However, six out of the 13 dropouts were due to death and seven were due to an increase in the severity of the diseases, which prevented them from going to the hospital for the intervention.

Behavioural approaches generally require an engagement of the participants in the intervention and cannot rely on strict double-blinding procedures.[70] Whether administrators or participants were aware of the intervention being administered may influence the outcome measure. For this reason, Domain 4 (risk of bias in measurement of the outcome) was rated as a medium risk of bias for all studies. Finally, for 10/11 studies, the risk of bias for the selection of the reported result (Domain 5) was low. The only medium risk was coded for one study where there was no description of planned analyses.

#### Acceptability, Feasibility, Fidelity of the Interventions

One objective of the study was to analyse the feasibility, acceptability, and fidelity of the interventions. The results will be presented according to each intervention program. More details can be found in the online supplemental eTable 2.

#### Hypnosis Interventions

All three hypnosis studies detailed their intervention. The intervention was always delivered by a hypnotherapist or therapists trained in hypnosis but only one[57] appears to have an intervention manual allowing for accurate replication of the procedure.

These studies also encountered some difficulties in terms of acceptability and feasibility. Indeed, two studies[51 54] observed a high attrition rate of their participants, representing respectively 26% and 48% (year 2). However, this dropout rate was largely explained by the death rate and by the deterioration of health. The evaluation of the participants’ interest in one study [51] nevertheless indicated that they found the intervention enjoyable and useful.

#### Hypnosis/Music Intervention

Both music/hypnosis interventions have a detailed intervention script. One study[50] followed all the steps for the development and validation of a complex intervention suggested by the UK Medical Research Council. The number of sessions (one or two) and the duration of each session (between 13 and 20 min) are concise. The other study[55] requires the presence of a music therapist and specific musical instruments. Finally, the attrition rate for both studies was also low, however, recruitment seems to have been challenging (e.g. 200 out of 400 patients agreed to take part in one study).[55]

#### Preferred Music

Interventions using preferred music are presented in three articles, however, none of them seem to follow a specific intervention manual. The intervention took place over one session for two studies[18 19] and over two sessions for the third.[53] Feasibility and acceptability are appropriate considering the low attrition rate observed.

#### Musicotherapy

For Musicotherapy Interventions, the three studies of this review provided a detailed protocol. For each of them, the interventions were detailed in the articles, although no manuals were identified. These interventions used several techniques that required the presence of a music therapist and specific equipment. The feasibility of the interventions was adequate, except for one intervention,[52] which used vibroacoustic stimulation, where the attrition rate was 44.4%. The authors also mentioned difficulties in recruiting participants. Nevertheless, patient satisfaction with the music therapy intervention, as assessed by one study[56] reached 95.6%.

## Discussion

This meta-analysis aimed to identify and evaluate the feasibility, acceptability, and protocol fidelity of hypnosis and music interventions in the context of PC, and the effects of these interventions on users. Through this review, we also aimed to assess whether the literature was sufficient to make informed choices about the parameters of music and hypnotic interventions that should be used for the management of pain, anxiety, sleep, and well-being in PC.

### Effect Sizes

The meta-analysis of RCT data from four studies shows a moderate and significant effect size on pain management in PC. This result combines all three types of intervention (hypnosis, music with hypnosis and music only). With these initial data, one may assume that these interventions may be appropriate for pain management in PC. However, this effect was primarily driven by the study of Gutgsell et al.[55] testing a combined intervention involving hypnosis and music in a large group of participants.

The pre-post data results of the 11 experimental groups allow us to consider more studies although they do not control for non-specific factors, such as the effect of the time and repetition of measures. However, we can observe with these preliminary results that the effect sizes of the hypnosis and preferred music interventions are very high for all four variables (pain, anxiety, sleep, and well-being) and three variables (pain, anxiety, and sleep). The effect is also high for music/hypnosis interventions for pain variable.

Considering the presence of large confidence intervals, indicating high variability in the results and/or small samples, we expect that by increasing the number of studies and participants, statistical power could be increased. There are very few studies evaluating hypnosis, music/hypnosis and music interventions for managing pain, anxiety, sleep, and well-being in PC, and even less study with an RCT design. We can conclude that the early studies tend to be favourable on the variables studied, however, it is too early to identify the characteristics of the interventions that play a key role in their effectiveness and to determine what type of intervention is most effective.

### Risk of Bias

The risk of bias varies across studies. However, the very nature of hypnosis and music interventions with people in PC make some criteria difficult to achieve.

First, given the difficulty of recruiting participants in the last moments of life and the ethical importance of providing the best care to individuals, it may be difficult to include a control group in the studies.

Secondly, for hypnosis or music RCT interventions, it is almost impossible to hide the nature of the intervention from participants and practitioners. Biases associated with beliefs about the effect of the intervention are therefore possible. Nevertheless, some authors have looked at research designs where it is not possible to have true double-blind procedure. These authors have suggested using a control condition of equivalent duration to the experimental condition for which the providers have an equivalent level of expertise. They also suggested that participants should be blinded to the intervention group (experimental or control).[70] The presence of a control group with characteristics as similar as possible to the experimental group could thus help reduce biases associated with belief in the intervention they belong to. Another strategy could be to assess participants’ level of confidence in the intervention before starting the experiment and to use this variable as a moderator.

Although some options exist to reduce the risk of bias, the quality assessment of PC studies involving behavioural interventions must be interpreted with caution as some criteria appear to be less appropriate in this clinical setting.

### Feasibility, acceptability, and fidelity

The feasibility analyses of the interventions provide a different perspective. Studies with multiple intervention sessions faced challenges in retaining participants. Considering that mortality and the deterioration of the health are inherent to end of life PC setting, we believe that particular attention should be paid to the duration of the interventions and the length of each session to ensure that benefits can be achieved rapidly. The intervention itself should be designed to minimise fatigue in participants.

The user’s interest in participating in the intervention is also an important aspect. By taking a more pragmatic approach and allowing participants to choose between different interventions, we can expect retention rates to be higher.

Particular attention should be paid to the development and use of a detailed intervention manual to ensure fidelity and consistency in the intervention. Few studies have reported using such a protocol, leaving room for significant variation in the intervention, minimising its fidelity and the validity of the study. However, a balance must be struck between achieving internal validity in “explanatory” studies and achieving external validity in pragmatic studies.[71] To ensure both the proper integration of an intervention in a setting and good internal validity, we believe that a detailed protocol that leaves room for a certain amount of flexibility in its implementation is a preferred option. This balance may be even more adapted to behavioural interventions intended to provide comfort care based on person-centred principles,[72] leaving the possibility of adapting the intervention to each user and to each environment.

### Limitations

The current study has some limitations. We only included articles in English and French. To achieve the objectives of this systematic review, we also excluded some studies because of the lack of precision of their protocol, thus reducing the size of our sample and the scope of the conclusions we can make. This limitation reflects the state of the field and strongly calls for improvement in the detailed description of the interventions in future studies.

### Future Directions

Hypnosis and music interventions for the management of pain, anxiety, sleep, and well-being in the PC setting are fairly recent. Nevertheless, the promising preliminary results obtained in the meta-analysis warrant the development of such interventions.

Future studies, with well-structured approaches, will undoubtedly highlight the essential components to be integrated into the interventions, their appropriate combination, as well as the length and number of sessions to be targeted.

For hypnosis interventions, research should focus on optimizing induction and suggestions in clinical contexts. With regard to musical interventions, the type of music to be used, the ideal duration, and how to integrate it into PC interventions are also variables to be further evaluated. Finally, although preliminary findings appear promising, the analysis of the added value of combining music and hypnosis interventions, as well as how to potentiate their respective effects, remains to be done more extensively.

### Conclusion

Despite the limitations associated with the small number of studies, the very large effect sizes obtained in several studies is stimulating and justify further studies of hypnosis, hypnosis-music, and music interventions for pain, anxiety, sleep, and well-being management in PC. These interventions have the potential to address an important need while presenting minimal risk of side effects. These approaches should be designed following the principles of person-centred medicine, taking into account individual preferences, yet this flexibility must be operationalised in intervention manuals to insure replicability.

## Supporting information

Supplemental eTable1

Supplemental eTable 2

Supplemental eFigure 1

## Data Availability

The data are from published studies that have been cited. Further details on the data analysis process can be provided on request.

## Acknowledgments

The authors would like to thank Kyle Roerick for its linguistic revision and Mélissa Gravel for her help in developing a search strategy.

## References

1. World B. Population âgée de 65 et plus (% du total) – North America, European Union 2021 [Available from: https://donnees.banquemondiale.org/indicateur/SP.POP.65UP.TO.ZS?end=2020&locations=XU-EU&start=1960&view=chart.

2. Organization WH. Palliative Care 2020 [Available from: https://www.who.int/news-room/fact-sheets/detail/palliative-care.

3. Cohen SR, Mount BM. Quality of life in terminal illness: defining and measuring subjective well-being in the dying. Journal of palliative care 1992;8(3):40–45.

4. Landry M, Stendel M, Landry M, et al. Hypnosis in palliative care: from clinical insights to the science of self-regulation. Annals of palliative medicine 2018;7(1):125–35.

5. Archie P, Bruera E, Cohen L. Music-based interventions in palliative cancer care: a review of quantitative studies and neurobiological literature. Supportive Care in Cancer 2013;21(9):2609–24. doi: 10.1007/s00520-013-1841-4

6. Parncutt R, McPherson G. The science and psychology of music performance: Creative strategies for teaching and learning: Oxford University Press 2002.

7. Adachi T, Fujino H, Nakae A, et al. A meta-analysis of hypnosis for chronic pain problems: a comparison between hypnosis, standard care, and other psychological interventions. International Journal of Clinical and Experimental Hypnosis 2014;62(1):1–28.

8. Chen PY, Liu YM, Chen ML. The effect of hypnosis on anxiety in patients with cancer: A meta-analysis. Worldviews on Evidence-Based Nursing 2017;14(3):223–36.

9. Lam T-H, Chung K-F, Yeung W-F, et al. Hypnotherapy for insomnia: a systematic review and meta-analysis of randomized controlled trials. Complementary therapies in medicine 2015;23(5):719–32.

10. Billot M, Jaglin P, Rainville P, et al. Hypnosis Program Effectiveness in a 12-week Home Care Intervention To Manage Chronic Pain in Elderly Women: A Pilot Trial. Clinical therapeutics 2020;42(1):221–29.

11. Dumain M, Jaglin P, Wood C, et al. Long-Term Efficacy of a Home-Care Hypnosis Program in Elderly Persons Suffering From Chronic Pain: A 12-Month Follow-Up. Pain Management Nursing 2021

12. MacDonald RA. Music, health, and well-being: A review. International journal of qualitative studies on health and well-being 2013;8(1):20635.

13. Chanda ML, Levitin DJ. The neurochemistry of music. Trends in cognitive sciences 2013;17(4):179–93.

14. Martin-Saavedra JS, Vergara-Mendez LD, Pradilla I, et al. Standardizing music characteristics for the management of pain: A systematic review and meta-analysis of clinical trials. Complementary therapies in medicine 2018;41:81–89.

15. Panteleeva Y, Ceschi G, Glowinski D, et al. Music for anxiety? Meta-analysis of anxiety reduction in non-clinical samples. Psychology of Music 2018;46(4):473–87.

16. Lu G, Jia R, Liang D, et al. Effects of music therapy on anxiety: A meta-analysis of randomized controlled trials. Psychiatry Research 2021;304:114137. doi: https://doi.org/10.1016/j.psychres.2021.114137

17. Gao Y, Wei Y, Yang W, et al. The effectiveness of music therapy for terminally ill patients: a meta-analysis and systematic review. Journal of pain and symptom management 2019;57(2):319–29.

18. Peng CS, Baxter K, Lally KM. Music Intervention as a Tool in Improving Patient Experience in Palliative Care. American Journal of Hospice & Palliative Medicine 2019;36(1):45–49.

19. Nakayama H, Kikuta F, Takeda H. A pilot study on effectiveness of music therapy in hospice in Japan. Journal of Music Therapy 2009;46(2):160–72.

20. Frohne-Hagemann I, Warja M, Pedersen IN, et al. Guided Imagery & Music (GIM) and Music Imagery Methods for Individual and Group Therapy. London: Jessica Kingsley Publishers, 2015.

21. McKinney CH, Honig TJ. Health Outcomes of a Series of Bonny Method of Guided Imagery and Music Sessions: A Systematic Review. Journal of Music Therapy 2016:thw016. doi: 10.1093/jmt/thw016

22. Melzack R. Pain–an overview. Acta Anaesthesiologica Scandinavica 1999;43(9):880–84.

23. Geuter S, Reynolds Losin EA, Roy M, et al. Multiple Brain Networks Mediating Stimulus-Pain Relationships in Humans. Cereb Cortex 2020;30(7):4204–19. doi: 10.1093/cercor/bhaa048

24. Wager TD, Atlas LY, Lindquist MA, et al. An fMRI-based neurologic signature of physical pain. N Engl J Med 2013;368(15):1388–97. doi: 10.1056/NEJMoa1204471

25. Bushnell MC, Ceko M, Low LA. Cognitive and emotional control of pain and its disruption in chronic pain. Nature reviews Neuroscience 2013;14(7):502–11. doi: 10.1038/nrn3516 [published Online First: 2013/05/31]

26. Tracey I. Neuroimaging of pain mechanisms. Current opinion in supportive and palliative care 2007;1(2):109–16. doi: 10.1097/SPC.0b013e3282efc58b [published Online First: 2008/08/08]

27. Juslin PN. From everyday emotions to aesthetic emotions: towards a unified theory of musical emotions. Physics of life reviews 2013;10(3):235–66. doi: 10.1016/j.plrev.2013.05.008

28. Mitchell LA, MacDonald RA. An experimental investigation of the effects of preferred and relaxing music listening on pain perception. Journal of music therapy 2006;43(4):295–316.

29. Roy M, Peretz I, Rainville P. Emotional valence contributes to music-induced analgesia. Pain 2008;134(1-2):140–47.

30. Roy M, Lebuis A, Hugueville L, et al. Spinal modulation of nociception by music. European journal of pain (London, England) 2012;16(6):870–7. doi: 10.1002/j.1532-2149.2011.00030.x [published Online First: 2012/02/18]

31. Pelletier CL. The effect of music on decreasing arousal due to stress: A meta-analysis. Journal of music therapy 2004;41(3):192–214.

32. Landry M, Lifshitz M, Raz A. Brain correlates of hypnosis: A systematic review and meta-analytic exploration. Neuroscience and biobehavioral reviews 2017;81(Pt A):75–98. doi: 10.1016/j.neubiorev.2017.02.020 [published Online First: 2017/02/28]

33. Rainville P, Duncan GH, Price DD, et al. Pain affect encoded in human anterior cingulate but not somatosensory cortex. Science (New York, NY) 1997;277(5328):968–71. doi: 10.1126/science.277.5328.968 [published Online First: 1997/08/15]

34. Hofbauer RK, Rainville P, Duncan GH, et al. Cortical representation of the sensory dimension of pain. Journal of neurophysiology 2001;86(1):402–11. doi: 10.1152/jn.2001.86.1.402 [published Online First: 2001/06/30]

35. Hilgard ER. A quantitative study of pain and its reduction through hypnotic suggestion. Proceedings of the National Academy of Sciences of the United States of America 1967;57(6):1581–6. doi: 10.1073/pnas.57.6.1581 [published Online First: 1967/06/01]

36. Hilgard ER, Morgan AH, Lange AF, et al. Heart rate changes in pain and hypnosis. Psychophysiology 1974;11(6):692–702. doi: 10.1111/j.1469-8986.1974.tb01138.x [published Online First: 1974/11/01]

37. Rainville P, Carrier B, Hofbauer RK, et al. Dissociation of sensory and affective dimensions of pain using hypnotic modulation. Pain 1999;82(2):159–71. doi: 10.1016/s0304-3959(99)00048-2 [published Online First: 1999/09/01]

38. De Pascalis V, Magurano MR, Bellusci A, et al. Somatosensory event-related potential and autonomic activity to varying pain reduction cognitive strategies in hypnosis. Clinical neurophysiology : official journal of the International Federation of Clinical Neurophysiology 2001;112(8):1475–85. doi: 10.1016/s1388-2457(01)00586-7 [published Online First: 2001/07/19]

39. Kiernan BD, Dane JR, Phillips LH, et al. Hypnotic analgesia reduces R-III nociceptive reflex: further evidence concerning the multifactorial nature of hypnotic analgesia. Pain 1995;60(1):39–47. doi: 10.1016/0304-3959(94)00134-z [published Online First: 1995/01/01]

40. Danziger N, Fournier E, Bouhassira D, et al. Different strategies of modulation can be operative during hypnotic analgesia: a neurophysiological study. Pain 1998;75(1):85–92. doi: 10.1016/s0304-3959(97)00208-x [published Online First: 1998/04/16]

41. Page MJ, McKenzie JE, Bossuyt PM, et al. The PRISMA 2020 statement: an updated guideline for reporting systematic reviews. BMJ 2021;372:n71. doi: 10.1136/bmj.n71

42. Higgins JP, Savović J, Page MJ, et al. Assessing risk of bias in a randomized trial. Cochrane handbook for systematic reviews of interventions 2019:205–28.

43. Linton M-J, Dieppe P, Medina-Lara A. Review of 99 self-report measures for assessing well-being in adults: exploring dimensions of well-being and developments over time. BMJ open 2016;6(7):e010641.

44. Ouzzani M, Hammady H, Fedorowicz Z, et al. Rayyan—a web and mobile app for systematic reviews. Systematic reviews 2016;5(1):1–10.

45. Czajkowski SM, Powell LH, Adler N, et al. From ideas to efficacy: The ORBIT model for developing behavioral treatments for chronic diseases. Health psychology : official journal of the Division of Health Psychology, American Psychological Association 2015;34(10):971–82. doi: 10.1037/hea0000161

46. Borenstein M, Hedges LV, Higgins JP, et al. Comprehensive meta-analysis (Version 2.2. 027)[Computer software]. Englewood, NJ: Biostat 2006

47. Morris SB. Estimating effect sizes from pretest-posttest-control group designs. Organizational research methods 2008;11(2):364–86.

48. Morris SB, DeShon RP. Combining effect size estimates in meta-analysis with repeated measures and independent-groups designs. Psychological methods 2002;7(1):105.

49. Duval S, Tweedie R. Trim and fill: a simple funnel-plot–based method of testing and adjusting for publication bias in meta-analysis. Biometrics 2000;56(2):455–63.

50. Coelho A, Parola V, gren A, et al. The Effects of Guided Imagery on Comfort in Palliative Care. Journal of Hospice & Palliative Nursing 2018;20(4):392–99.

51. Plaskota M, Lucas C, Evans R, et al. A hypnotherapy intervention for the treatment of anxiety in patients with cancer receiving palliative care. International Journal of Palliative Nursing 2012;18(2):69–75.

52. Warth M, Kessler J, Kotz S, et al. Effects of vibroacoustic stimulation in music therapy for palliative care patients: a feasibility study. BMC Complementary & Alternative Medicine 2015;15(1):436.

53. Warth M, Kessler J, van Kampen J, et al. ‘Song of Life’: music therapy in terminally ill patients with cancer. BMJ supportive & palliative care 2018;8(2):167–70.

54. Brugnoli MP, Pesce G, Pasin E, et al. The role of clinical hypnosis and self-hypnosis to relief pain and anxiety in severe chronic diseases in palliative care: a 2-year long-term follow-up of treatment in a nonrandomized clinical trial. Annals of Palliative Medicine 2018;7(1):17–31.

55. Gutgsell KJ, Schluchter M, Margevicius S, et al. Music therapy reduces pain in palliative care patients: a randomized controlled trial. Journal of Pain & Symptom Management 2013;45(5):822–31.

56. Planas-Domingo J, Escudé Matamoros N, Farriols Danés C, et al. Effectiveness of Music Therapy in Advanced Cancer Patients Admitted to a Palliative Care Unit: A Non-Randomized Controlled, Clinical Trial. Music & Medicine 2015;7(1):23–31.

57. Liossi C, White P. Efficacy of clinical hypnosis in the enhancement of quality of life of terminally ill cancer patients. Contemporary Hypnosis (John Wiley & Sons, Inc) 2001;18(3):145–60.

58. Warth M, Kesler J, Hillecke TK, et al. Music Therapy in Palliative Care. Deutsches Arzteblatt International 2015;112(46):788–94.

59. Hamilton M. The assessment of anxiety states by rating. British journal of medical psychology 1959;32(1):50–55.

60. De Haes J, Olschewski M, Fayers P, et al. The Rotterdam symptom checklist (RSCL). Amsterdão: University of Groningen 1996

61. Zigmond AS, Snaith RP. The hospital anxiety and depression scale. Acta psychiatrica scandinavica 1983;67(6):361–70.

62. Snyder-Halpern R, Verran JA. Instrumentation to describe subjective sleep characteristics in healthy subjects. Research in Nursing & Health 1987;10(3):155–63.

63. Nekolaichuk C, Watanabe S, Beaumont C. The Edmonton Symptom Assessment System: a 15-year retrospective review of validation studies (1991–2006). Palliative medicine 2008;22(2):111–22.

64. Voepel-Lewis T, Shayevitz JR, Malviya S. The FLACC: a behavioral scale for scoring postoperative pain in young children. Pediatr Nurs 1997;23(3):293–97.

65. Gloth III F, Scheve A, Stober C, et al. The Functional Pain Scale: reliability, validity, and responsiveness in an elderly population. Journal of the American Medical Directors Association 2001;2(3):110–14.

66. Caraceni A, Cherny N, Fainsinger R, et al. Pain measurement tools and methods in clinical research in palliative care: recommendations of an Expert Working Group of the European Association of Palliative Care. Journal of pain and symptom management 2002;23(3):239–55.

67. Hara N. Kibun chosa-hyou [Mood inventory]. In H. Hori & M. Yamamoto (Eds.), Shinri-sokutei shakudo shu I: Ningen no naimen o saguru [A collection of psychological scaling measures: Vol. 1. Exploring inner beings of people] (pp. 249–255). Tokyo: Science-sha. 1994

68. Chiu L, Chiu N, Chow E, et al. Comparison of three shortened questionnaires for assessment of quality of life in advanced cancer. Journal of palliative medicine 2014;17(8):918–23.

69. Borenstein M, Hedges LV, Higgins JP, et al. Introduction to meta-analysis: John Wiley & Sons 2021.

70. Davidson RJ, Kaszniak AW. Conceptual and methodological issues in research on mindfulness and meditation. Am Psychol 2015;70(7):581–92. doi: 10.1037/a0039512

71. Patsopoulos NA. A pragmatic view on pragmatic trials. Dialogues Clin Neurosci 2011;13(2):217–24. doi: 10.31887/DCNS.2011.13.2/npatsopoulos

72. Mezzich JE, Kirisci L, Salloum I, et al. Systematic conceptualization of person centered medicine and development and validation of a person-centered care index. International Journal of Person Centered Medicine 2016;6(4)

