## Supplemental eTable1 for "Hypnosis and music interventions for pain, anxiety, sleep, and well-being in palliative care: a systematic review and meta-analysis"

RESEARCH STRATEGY

Concepts and terms used:

INTERVENTIONS

Concept 1 – hypnosis or hypnotherapy or hypnoses or hypnotism or hypnotherapies or "guided imagery" or sophrology

Concept 2 – Music*

OUTCOMES

Concept 3 – Pain

Concept 4 – Anxiety or fear or "fear of dying " or "fear of death " or "psychological stress" or distress

Concept 5 – Sleep or insomnia

Concept 6 – Well-being or "psychological state" or "Quality of life" or "Life satisfaction"

CLINICAL SETTINGS

Concept 7 – "Palliative care" or "end of life care" or "terminal care" or dying or "hospice care"

Databases and interfaces:

| Interface | Database | Categories |
| --- | --- | --- |
| Embase | Embase | Title, Abstract, OR Keyword |
| Ovid | PsychINFO; PubMed | Abstract, Heading word, Keyword heading word OR Title |
| Ebsco | CINAHL Plus  CINAHL  Psychology and Behavioral Sciences Collection  AgeLine | Title, Subject OR Abstract |

Steps of research:

1) Search for single concepts

2) Combining concepts for documentary research

(Concept 1 OR Concept 2) AND (Concept 3 OR Concept 4 OR Concept 5 OR Concept 6) AND (Concept 7)

Limits:

- English and French
- Peer reviewed
- Human
- Clinical trial
