## Supplemental eTable 2 for "Hypnosis and music interventions for pain, anxiety, sleep, and well-being in palliative care: a systematic review and meta-analysis"

*eTable 2. Description of the interventions*

**HYPNOSIS**

| Author | Aim of the study | Theoretical models explaining needs and difficulties by patient in PC | Theoretical models explaining impact of intervention | Models of change | | Concrete intervention techniques implemented | Criteria study | | |
| --- | --- | --- | --- | --- | --- | --- | --- | --- | --- |
|  |  |  |  |  |  |  | Acceptability | Feasibility | Fidelity |
| Brugnoli et al. (2018) | “To investigate the efficacy of hypnosis and self-hypnosis in relieving pain and anxiety, in severe chronic diseases for a long-term follow up of 1 and 2 years.” | “Chronic pain conditioning [as a] form of cognitive learning in which subjects come to express responses to neutral conditioned stimuli, which are paired with aversive unconditioned stimuli.  “Because of this pairing, the CS acquires the ability to elicit a spectrum of behavioral, autonomic, and endocrine responses that would normally only occur in the context of danger” ^70^  “Chronic pain conditioning can be adaptive and enable efficient behavior in situations like severe chronic diseases”.  “The long loop pathway indicates that sensory information relayed to the amygdale undergoes substantial higher level processing, thereby enabling the assignment of significance, based upon prior experience of anxiety, to complex stimuli.”(Blair, 2001) | Hypnosis “focus on the interactions between the brain, body, mind, spirit and behavior, with the intent of using the mind to modify physical function and behaviors and promote overall physical (pain) and psychological (anxiety) health”.  “Hypnosis modulates phenomenological aspects of conscious experience, such as pain perception”.  “Hypnosis influences anxiety symptoms through emotional regulatory mechanisms by considering emotional regulation as a path through which clinical hypnosis stimulates mental health” (Erikson, 1980).  “Hypnotic analgesia arguably originates from various factors, including the alteration of expectations relative to impending painful events, as well as attentional, cognitive and emotional regulation mechanisms”. | | “To teach the hypnosis group patients clinical hypnosis and self-hypnosis as an adjuvant therapy to their pharmacological therapy in order to relieve pain and anxiety”. | “2-hours workshop with PI, which explained the meaning of clinical hypnosis in palliative care for pain and anxiety relief”.  “For 2 years, a series of weekly, 2-hour workshops on chronic pain assessment and the management of anxiety and related symptoms”.  “Teaching the hypnosis group patients how to use hypnosis and self-hypnotic techniques [...] The patients could decide whether to use just one or several of the preferred self hypnosis techniques at home.  Techniques of clinical hypnosis and self-hypnosis used:  *“-The different interpretation of symptoms technique*  *-Example of anesthesia in one hand*  *-Exercise “warm hands” (or for different parts of the body)*  *-The transferred symptoms technique*  *-The positive visualization technique (for pain and anxiety relief) (safe place)*  *-The desensitization of pain and anxiety technique*  *-Self-hypnosis technique for pain and anxiety relief*  *-The self-hypnosis CD method”* | After 1 year, no dropouts.  13 dropouts out of 50 at year 2 (26%).  No side effects noted. | Difficulty of recruitment.  Possible to do a 2 years intervention. | Description of the intervention but no manual mentioned for each of the 104 interventions. |

| Author | Aim of the study | Theoretical models explaining needs and difficulties by patient in PC | Theoretical models explaining impact of intervention | Models of change | Concrete intervention techniques implemented | Criteria study | | |
| --- | --- | --- | --- | --- | --- | --- | --- | --- |
|  |  |  |  |  |  | Acceptability | Feasibility | Fidelity |
| Liossi and White (2001) | “To assess the efficacy of clinical hypnosis in the management of depression and anxiety, and the enhancement of quality of life for patients with far-advanced cancer”. | “Terminal cancer patients experience considerable psychological distress” (Breitbart and Passik, 1993).  “Anxiety, demoralization, suffering, isolation, anger and depression are especially relevant to patients with advanced cancer”.  “Dying patients experience a heavy physical symptom burden”.  Psychological distress can occur “as a consequence of specific negative conditions inherent in the disease, such as  - pain and other symptoms, ambiguity, conflict, novelty and complexity :  -one’s appraisal of the disease as a threat to the physical, psychological, spiritual and social existence of the individual;  -a poor, self-assessment of one’s ability to cope with the threat; -ineffective attempts at problem-solving and finding meaning in the life lived and beyond the lifespan”. | “Clinical hypnosis has been used successfully for the alleviation of a variety of cancer-related symptoms, including pain”.  “It promotes a sense of control in the patient over the effects of cancer and can easily be combined with and enhance the effect of other psychotherapeutic modalities”.  Allow to work with “patients to provide them with a skill with which they can manage their symptoms and gain a sense of self-worth and self-efficacy”. | To focusing on “symptom management and ego-strengthening”.  To reducing emotional distress (“anxious mood, restlessness and anxious thoughts, depression, grief, demoralization, low self-esteem and pessimism”).  -To “improving mental adjustment to cancer “.  -To “promoting effective coping strategies”. | The hypnosis intervention (4 x 30 min) consisted of psychological support,” induction, suggestions for symptom management and ego strengthening, and post-hypnotic suggestions for comfort and maintenance of the therapeutic benefits during the following week”.  -“Inductions used were the arm levitation and the cloud fantasy technique. Relevant suggestions were made according to patients’ predominant symptoms and requests […] (ex. analgesic suggestions for residual pain and suggestions for diminished anxiety, for nausea and vomiting management, insomnia, breathlessness and fatigue).  “Ego-strengthening consisted of general ego-strengthening suggestions, specific ego-strengthening suggestions to facilitate the discovery and enhancement of patients’ inner coping strategies and specific suggestions to foster patients’ sense of self-efficacy (Brown and Fromm, 1986)”.  Standard care  “The psychological support consisted of supportive counselling based on the cognitive existential therapeutic tradition”. | Low attrition rate.  Semi-structured interviews: Satisfaction seems good. | Yes | Treatment manual was prepared. |

| Author | Aim of the study | Theoretical models explaining needs and difficulties by patient in PC | Theoretical models explaining impact of intervention | Models of change | Concrete intervention techniques implemented | Criteria study | | | |
| --- | --- | --- | --- | --- | --- | --- | --- | --- | --- |
|  |  |  |  |  |  | Acceptability | Feasibility | | Fidelity |
| Plaskota et al. (2012) | “To assess the effectiveness of hypnotherapy in the management of anxiety in palliative care patients”.  “To ascertain whether hypnotherapy could enhance sleep quality and quantity, reduce frequency of sleep disturbances, and reduce the severity of cancer-related depression and other psychological and physical symptoms”. | “Anxiety is more common in patients with cancer than in healthy populations (Jackson and Lipman, 2000)”.  “In palliative care patients, anxiety is often attributed to a reaction to diagnosis, treatment, and its possible outcomes”.  “Fears about impending death may well be present, but they are not necessarily the main source of anxiety in terminally ill patients”.  “Concerns about family and loved ones may contribute (Jackson and Lipman, 2000), and several organic causes may also co-exist”. | “Hypnotic inductions closely resemble these conventional relaxation and psychotherapy techniques and may be a vehicle by which these techniques can be taught (Kirsch et al, 1995; Douglas, 1999)”.  “Liossi and Mystakidou (1996) describe hypnosis as a psychological state that may serve to heighten some human capacities while allowing others to fade into the background”.  “It has the benefit of being a non-invasive intervention free from pharmacological side effects”. | To manage of anxiety in palliative care patients “by an induction procedure that includes suggestions of relaxation, calmness, and wellbeing”. | Beginning of each session: “discussion with the patient about their concerns and how hypnotherapy treatment might help him/her and then continued as outlined below”.  First session - self-hypnosis: “At each session, induction of hypnotherapy trance was achieved using eye fixation and progressive relaxation. In trance, the patient was taught how to access and use positive self-suggestion”.  Second session - visualization: “This technique involved helping the patient to perceive a troubling symptom as an external entity and to manage it as if it were a physical reality. Intrusive and negative thoughts may also have been addressed through visualization”.  Third session- install an anchor: “The patient was taught to use an ‘anchor’ to access a preferred mental state. This involved recalling a positive memory and linking this with a physical trigger, e.g. a finger pinch. The physical trigger was then used by the patient at will when they were faced with a difficult situation or emotion”.  Fourth session: immune system visualization: “This process aimed to enable the patient to visualize his/her immune system in a dynamic way, in the hope of consciously enhancing the immune process. It is a tool to enhance the individual’s sense of control over their disease process. All patients were brought out of trance in a controlled manner and reorientated to their current environment. The order of these techniques may have been altered depending on the patient’s particular issues and needs”. | High attrition rate: 2 deaths, 5 due to physical deterioration and 3 patients voluntarily withdrew from the study.  Difficulties of recruitment. | | Satisfaction survey positive: five of the eleven patients found the hypnotherapy sessions very enjoyable; found them enjoyable.  Eight of the patients found them very helpful and the remaining three found them helpful. | Outline of the intervention is written. No manual mentioned. |

Music/ Hypnosis

| Author | Aim of the study | Theoretical models explaining needs and difficulties by patient in PC | Theoretical models explaining impact of intervention | Models of change | Concrete intervention techniques implemented | Criteria study | |
| --- | --- | --- | --- | --- | --- | --- | --- |
|  |  |  |  |  |  | Acceptability and  Feasibility | Fidelity |
| Coelho et al. 2018 | To evaluate the effects of GI on the comfort, pain, heart rate, and respiratory rate of patients in PC. | PCU patients :  -“Basic needs for relief, tranquility, and transcendence” (Kolcaba, 2003)  -“Need to be fortified for difficult tasks such as death”.  “Discomfort experienced by PCU patients was due to the loss of freedom and the absence of contact with an external green space”.  “The experience of comfort may emanate from dedicating time to relationships”. ^71^ | -To produce neurochemical changes with profound psychophysiological consequences by the intensity of a person’s involvement with mental images that allow his/her body to react as if responding to a genuine external experience.  -To enhances the effect of the mental images by music.  -To improve transcendence -through memorable connections between the nurse and patient.  -To “making the patients feel strengthened in intangible, personalized ways”. | | 2-session GI program.  “Prior to the first session, each patient chose the most comforting environment for him/her: a field or a beach”.  Intervention :  -“General indications that included the name of the technique and instructions on the attitude and posture to be adopted”;  -“Smooth breathing exercises and muscle relaxation, in an effort to eliminate some existing tension”;  - “induction of a sequential set of mental images that include special, quiet, and comfortable places enhancing the security and freedom of the patient”.  -Elaboration “by the patient on a sequential set of mental images, evoking natural scenarios in which to move, focusing particularly on the sensory content of these scenarios through vision, hearing, smell, and touch”.  -Suggestion to the patient “to move from this place and to take a walk, during which he/ she can imagine a meeting with someone special, allowing his/her a moment for meeting and sharing”.  -Invitation to the patient “to leave the place and to bring with him/her what felt good, leaving the state of relaxation”.  It was finally “suggested that the patient look at the PCU as a space where there are health care professionals who can help them feel comfortable”.  “Both sessions were accompanied by relaxing music concordant with the selected environment”. | Construction and validation of the adjusted GI program incorporated the guidelines and recommendations of the Medical Research Council (Craig et al., 2008) for the development of complex interventions (see Coelho, 2018).  Consensus among experts, nurses, and patients hospitalized in the PCU about the relevance of the intervention, the quantitative and qualitative evaluation during the field testing sub-phase has been obtained before experimentation.  Low attrition rate: 2/28 | Script for the intervention. |

| Author | Aim of the study | Theoretical models explaining needs and difficulties by patient in PC | Theoretical models explaining impact of intervention | Models of change | Concrete intervention techniques implemented | Criteria study | |
| --- | --- | --- | --- | --- | --- | --- | --- |
|  |  |  |  |  |  | Acceptability and feasibility | Fidelity |
| Gutgsell et al. (2013) | “To determine the efficacy of a single music therapy session to reduce pain in palliative care patients”. | “Medications that lower pain may lower patients’ sense of control and have unwanted side effects such as sedation, nausea, and constipation”.  “Patients and families may fear addiction to opioids. Pain medications primarily target the sensory (intensity) dimension of pain” (Kwekkeboom, 2008). | “To assist the patient in regaining self-control and becoming actively involved in the management of his/ her pain”.  (Use of AMTA fact sheet on pain management) | | -“Preparing the patient and the environment (adjusting the lights, offering a blanket, turning off cell phones, and so on, ‘‘Do Not Disturb’’ sign on the door”.  -Choice to the patient (ocean drum or not)  -“One 20-minute MT intervention directed at lowering pain” (standard protocol for all participants).   a. Verbal instructions for *autogenic relaxation*. The patient was ask to pay attention :   - “to breathing for approximately one minute”.  -“to the scalp muscles and allow them to release, and moving down with similar focus on specific muscle groups, ending with the feet”.   b. Safe place   -The patient was invited to imagine a safe place of his or her own choosing. “The therapist asked the patient to imagine what he or she saw, smelled, heard, tasted, and felt on the skin at the safe place.”   c. Music   -“MT informed the patient that she would begin to play first the ocean drum, if chosen, and then the harp to support his or her exploration of the safe place. The therapist played the same harp pieces for every patient. All pieces were played at a soft volume in a slow tempo and are described as follows:   - an improvisation in the mode of G Mixolydian with a duple meter, - four precomposed pieces in the key of C Major that can be described as ‘‘light classical’’ and are unfamiliar to most listeners”:‘  d. Conclusion   MT invited “the participant to leave his or her imagined safe place and re-enter the hospital room, realizing that the safe place is a resource to which he or she can return at any time. Then the music therapist left the room”.  The therapist “gave each study participant a CD of the intervention for future use and provided a CD player on request. Interested readers may contact the investigator to request a recording of the intervention”. | Attrition of experimental group: 1/100.  Recruitment: 200 out of 400 patients agreed to participate in the study. | Standard protocol. |

Music

Preferred music

| Author | Aim of the study | Theoretical models explaining needs and difficulties by patient in PC | Theoretical models explaining impact of intervention | Models of change | Concrete intervention techniques implemented | Criteria study | | |
| --- | --- | --- | --- | --- | --- | --- | --- | --- |
|  |  |  |  |  |  | Acceptability | Feasibility | Fidelity |
| Nakayama (2009) | To determining the effectiveness of music therapy in a hospice setting patients by measuring s-cortisol and levels changes in mood. | “With the advancement of medical technology, […] the survival rate of cancer patients has been markedly improved”. “There is a strong movement toward emphasis on QoL in addition to removal of lesions, and psychological and spiritual care in cancer treatment is called for”. | Music has an immediate impact on the psychological state, such as changing mood and evoking emotional reactions that are associated with past experience, by diverting attention from a certain stressor to music, or being a have positive stimulus (White, 2000).  Music is also suggested to physiological impact such as lowering blood pressure and heart rate and increasing the immunoglobulin A (IgA) level.  IgA is responsible for local immunity on mucous membranes against bacteria and viruses and known to decrease stress. That is, music may help improve immunity. | To Influence the psychological state and stress of the individual through music. | Context  “A weekly music session is usually held in the hospice air- conditioned multipurpose hall”.  Intervention  Few days before the experimentation, the nurses had asked them if they had any particulars requests for the session.  Focusing on their requests, songs that were appropriate for the season and the age-group were selected such that the number of the major and minor keys and slow-paced and up-tempo songs were as even as possible.  “Musical instruments used were piano, flute, little instruments for easy participation of the patients such as maracas, touch-bells, and the voice”.  “The song list included Hotaru Koi, which is one of Japanese traditional children's songs, Moldau, which was played by the piano intending harmonic and soft repetition of the rhythm, and Ue O Muite Aruko, which is a very popular song and was sang cheerfully”. | NA  Attrition | Participants regularly attend music therapy sessions | Outline of the intervention is written. No manual mentioned. |

| Author | Aim of the study | Theoretical models explaining needs and difficulties by patient in PC | Theoretical models explaining impact of intervention | Models of change | Concrete intervention techniques implemented | Criteria study | |
| --- | --- | --- | --- | --- | --- | --- | --- |
|  |  |  |  |  |  | Acceptability and feasibility | Fidelity |
| Peng et al. (2019) | “This pilot study aimed to examine palliative care patients’ symptomatology in response to receiving music intervention from behavioral, biological, and subjective experiential standpoints”. | “Patients in palliative care frequently struggle with symptom management”.  “The pain, anxiety, and stress associated with end-of-life care are paramount issues to address for both patients and their families”. | To “offering live music with a musician”. (“A live musician can interact with a patient and modify their music choice based on the patient’s wants and needs in a dynamic and intuitive fashion”.)  “To ensured individualized music selection that gave patients the ability to choose their preferred music”.  “To empowered patients, granting back the important impetus of control that many may have relinquished over the course of disease”.  “To use the music as a platform to reminisce about their lives or simply as a tool to reflect and relax”.  “To offers a social component to the experience”.  “The mechanism of music intervention can allow patients to be as proactive as they would like, with reactions of singing and clapping along to resting and reclining while just listening”. | | “The musician ascertained the patient’s musical preferences and performed the appropriate selections for the patient”.  “The musician who performed the intervention ad a variety of selections for the flute, ensuring a diverse assortment of musical genres to meet patients’ preferences. The genre and specific pieces of music were chosen in collaboration between the patient, the musician, and the family (if applicable”).  “Each encounter was idiosyncratic in that it served patients in a way that was most personally beneficial, be that an opportunity to use the music as a platform to reminisce about their lives or simply as a tool to reflect and relax”. | Positive experience by participants as mentioned in the interview.  Attrition : 3/46 | Outline of the intervention is written. No manual mentioned. |

| Author | Aim of the study | Theoretical models explaining needs and difficulties by patient in PC | Theoretical models explaining impact of intervention | Models of change | Concrete intervention techniques implemented | Criteria study | |
| --- | --- | --- | --- | --- | --- | --- | --- |
|  |  |  |  |  |  | Acceptability and  Feasibility | Fidelity |

| Warth et al. (2018)  Song of Life (SOL) | To assess “feasibility and acceptance of a novel MT technique called ‘Song of Life’ (SOL), targeting the improvement of emotional and spiritual well-being of terminally ill patients with cancer”. | “Patients prevalently show increased levels of psychological distress and a need for social and spiritual support”. | “Dignity therapy or other life review and meaning-centred techniques are examples of more recently emerging techniques specifically designed to address the needs of patients approaching death”. (Chochinov, 2011).  “Expression through music has shown to create feelings of meaning and hope for the patient and their families regarding coping with the anticipated threat of mortality and bereavement after death” (Magill, 2009; O'Callaghan, 2014).  “Songs were hypothesised to carry individual meaning and to be closely related to a person’s spiritual beliefs”. (Alvarenga, 2017) | -To” taking into account the characteristics of palliative care”.  -To “carry individual meaning and to be closely related to a person’s spiritual beliefs”.  -“To address the needs of patients approaching death by creating some sort of legacy based on the biographical history of the patient”. (Ando, 2010). | Session 1  Part 1: “Semi-structured interview on the patient’s life history, important relationships and roles”.  Part 2: Exploration of the patient’s SOL (“a song with a particularly high biographical relevance and the potential to arouse positive memories and strong emotional reactions in the listener”). “Questions guiding and supporting the patient in finding a SOL addressed relevant persons, situations, times and events that were related to a certain song (e.g. ‘Is there a song that reminds you of an important person/event/ place/location in your life?’). Patients were asked to choose a song which they perceived as a resource and not as a burden in that moment. In case the patient was unable to identify a personal SOL, the therapist suggested a popular song depending on the patient’s age.  Session 2 :  “The therapist used the guitar and her voice and first initiated a brief relaxation exercise accompanied by chords on the guitar in the musical key of the song. The music afterwards turned into a live performance of the SOL in a lullaby style. This required the therapist to translate the original version into a slow 3/4 or 6/8-rhythm, which showed to promote entrainment and relaxation in a previous study with newborn infants. The song was faded out with a diminuendo of the hummed chorus”.  “Afterwards, the therapist left the room for 10 min, giving the patient the opportunity to reflect on his/her experiences”.  “The therapist then returned for a debriefing conversation and for postintervention assessment of the LCS and VAS”. | Attrition : 2/15 | Outline of the intervention is written. No manual mentioned. |
| --- | --- | --- | --- | --- | --- | --- | --- |

| Musicotherapy | | | | | | | |
| --- | --- | --- | --- | --- | --- | --- | --- |
| Author | Aim of the study | Theoretical models explaining needs and difficulties by patient in PC | Theoretical models explaining impact of intervention | Models of change | Concrete intervention techniques implemented | Criteria study | |
|  |  |  |  |  |  | Acceptability and feasibility | Fidelity |
| Planas-Domingo et al. (2015) | “To analyze the effectiveness of music therapy interventions in advanced cancer patients admitted to a PCU on physical symptoms (pain and other), emotional symptoms (depression, anxiety) and well-being”. | “Advanced and terminal illness is associated with many physical and emotional symptoms, an emotional impact on the patient, family and health care team, and a limited survival prognosis”.  “In patients with terminal illness, fluidity of speech and discourse is often absent, arguably due to the uncertainty of their future, as well as physical limitations or inability to openly express their feelings. Verbal communication is often difficult due to the intensity of the emotions they are experiencing, or because of a desire to spare their loved ones”.  “Often, pharmacological treatment is not enough to improve emotional symptoms such as insomnia, depression, and anxiety”.  “The person who is dying highly appreciates the reaffirming proximity of others, active and attentive listening, and being valued for who he or she is and who he or she has been”. | “MT may contribute to each patient and families being able to explore opportunities to enjoy time together or in solitude, to review life and to achieve a sense of completion in relationships and life itself”.  “MT enables validation of feelings and releasing them. It enables the communication of important messages by a population with a strong need to convey wishes and desires". (Krout, 2003)  Music” may offer relief not only from the physical symptoms but also psychosocial, and spiritual problems in patients receiving palliative care”.  “Specific songs or works may hold special meaning through associative processes in one's life experience”.  MT “methods can improve self-esteem and provide to the person with an incentive and motivation to live fully throughout the process of dying”. | | Session 1 :  “Initiate contact with the patient and carry out the first data capture, it was common to also provide a music therapy intervention”.  Session 2 (day 3) :  “Played the songs that the patient had indicated he or she preferred during the gathering of the patient’s musical history. The patient sang or listened actively.  The lyrics were usually discussed and analyzed, or the feelings that the song evoked were processed.  According to prior assessment, the music therapists provided one or a number of the following interventions:  -music sedation  -guided visualization with music (guiding the patient to a significant life event accompanied by relaxing music played on guitar, ocean drum or clarinet);  -music assisted (inviting the patient to close their eyes using, gentle prompts in the nurturing presence of the therapist accompanied by music in minor mode on guitar, with the entraining music with the patient’s breathing).  Post session, the images and the patient’s comments that emerged during relaxation were processed. After, the patient would verbalize feelings, or communicate them non-verbally”.  Session 3 (day 5)  “Followed the same procedure as the second. Song writing was provided according to assessment”.  Session 4 (day 7)  “Family members were invited to the session to share the song created by the patient, along with his or her favorite song from their musical history. Experiences, emotions and memories were explored.”  Modality  “Live music using voice and various instruments (not recorded music”).  “Different personalized music interventions were used: singing, active music listening, song-writing, musical life review, playing an instrument, music sedation and visualization with music.  Instruments used were: guitar, ocean drum, violin, harp and tambura. Voice or, sometimes, another patient preferred instrument such as saxophone or clarinet were also used”. | 64 patients on 658 admitted to PCU were eligible and accepted to take part to the study.  Patient satisfaction with music therapy session was 3.8/4 for the 3 firsts sessions and 3.9/4 for the last one. | Outline of the intervention is written. No manual mentioned. |

| Mindfulness + music improvisation by MT | | | | | | | |
| --- | --- | --- | --- | --- | --- | --- | --- |
| Auteur | Aim of the study | Theoretical models explaining needs and difficulties by patient in PC | Theoretical models explaining impact of intervention | Models of change | Concrete intervention techniques implemented | Criteria study | |
|  |  |  |  |  |  | Acceptability  Feasibility | Fidelity |
| Warth et al. (2015a)  live music–based relaxation exercises | “This study examined whether relaxation interventions as part of music therapy could be successfully used to achieve the following endpoints:  1-Improvement in self-rated relaxation, well-being, and acute pain (primary endpoints)  2- Triggering of a physiological relaxation response 3- Improvement in health-related quality of life. It was expected that there would be improvements in  both study groups, and that music therapy would be shown to be superior”. | Patients with incurable illnesses have physical, psychosocial and spiritual needs.  Their quality of life is impaired (symptom, emotion, communication). |  | To support of “symptom management, improvement in regulation of emotions, and enhancement of communication and spiritual experiences. | Music Intervention   1) “Short mindfulness exercise, accompanied by soft monochord sounds. 2) Taking account of the patient’s breathing pattern, the volume, dynamics, and intensity of the monochord playing were then increased, and vocal improvisation was begun in Ionian or Mixolydian mode. 3) Towards the end of the improvisation, intensity was gradually reduced (approximately 15 minutes). 4) In a five-minute discussion after the procedure, the patient had the opportunity to reflect on his or her experience of listening to the music”. | Attrition 6/84  Recruitment 84/153 | Outline of the intervention is written. No manual mentioned. |

| Auteur | Aim of the study | Theoretical models explaining needs and difficulties by patient in PC | Theoretical models explaining impact of intervention | Models of change | Concrete intervention techniques implemented | Criteria study | |
| --- | --- | --- | --- | --- | --- | --- | --- |
|  |  |  |  |  |  | Acceptability and  feasibility | Fidelity |
| Warth et al. (2015 b) | “To investigate whether it was feasible to transfer elements of the previous research design to the evaluation of the effects of vibroacoustic stimulation for advanced cancer patients in palliative care”. | People in hospice care have physical symptoms: (e.g. pain, stress symptoms, shortness of breath), emotional and spiritual needs and communication issues. | “The use of vibroacoustic stimulation in medical settings is the idea that musical and emotional experiences may be intensified if a person not only listens to music, but actually senses the tactile vibration”.  “In contrast to active music therapy - where the patient himself plays an instrument or uses his voice – receptive techniques require very little or no physical activity and mental strain, and thus, are frequently used in end-of-life care settings”. | “To improve or maintain the patient’s quality of life by supporting physical symptom management (e.g. pain, stress symptoms, shortness of breath), addressing emotional needs, enhancing communication, and facilitating spiritual experiences”.  “To feel more relaxed and calm and to have experienced pleasant body sensations and visualizations after a music therapy session using the body tambura”. | “Short body scan (i.e. a mindfulness exercise focusing attention on different body parts) accompanied by slow and gentle play of the backside strings, lasting for approximately 3 min”.  “The therapist initiated a vocal improvisation in an Ionian or mixolydian mode while she gradually increased the volume, dynamics, and range of her musical play. During this improvisation, the music therapist tried to synchronize and modify musical and breathing meter”.  “After 10–12 min, the intensity of music and singing were slowly reduced and finally faded out. The therapist asked the participant to return his attention to the present moment”.  “The patient had the opportunity to reflect on his subjective experiences during the singing chair music”. | “In 5 out of 9 participants (56 %) the intervention and study protocol could be delivered as intended”.  Six semi-structured interviews were completed.  “Recruitment of participants was difficult and time-consuming. Many patients were not eligible for study participation due to the exclusion criteria.  Particularly, the requirement to sit in an upright position for up to half an hour led to a reduction in the number of eligible subjects”.  “Although most patients perceived the sounds and tactile stimulation as positive and calming, some patients reported experiences of arousal or being overwhelmed”.  Recruitment rate : 9/20 | Outline of the intervention is written. No manual mentioned. |
