## Supplementary figures and images for "Hypnosis and music interventions for pain, anxiety, sleep, and well-being in palliative care: a systematic review and meta-analysis"

### Supplemental eFigure 1

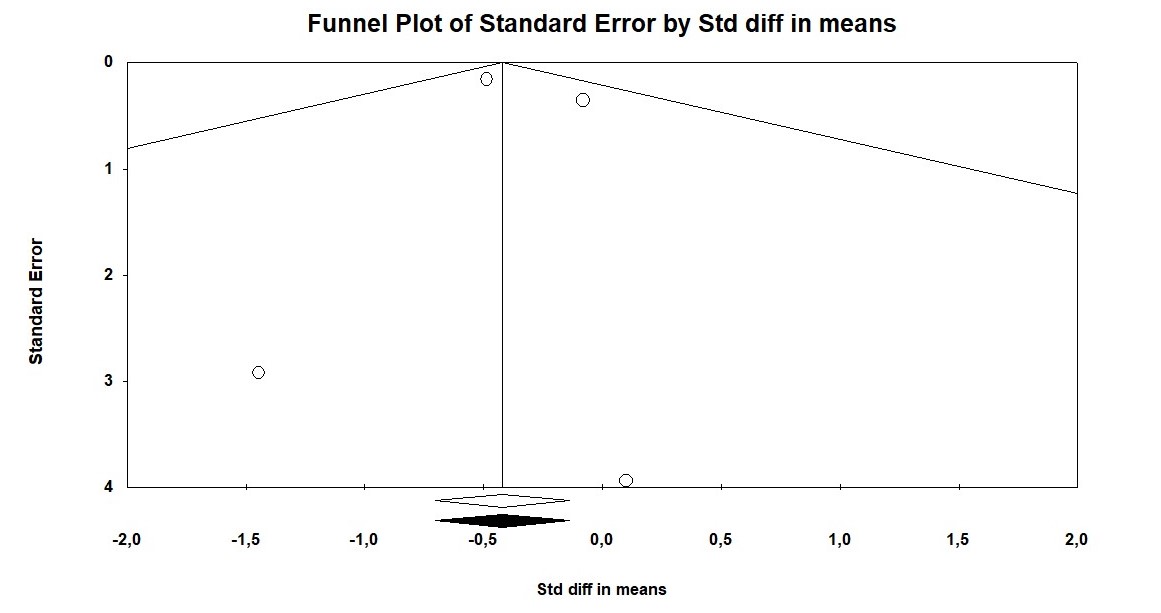
